# A minimal set of prompts to surface reasoning in digital health

**DOI:** 10.1101/2025.09.22.25336413

**Authors:** Ilona Kulikosvkikh, Tarzan Legović, Sergey Konovalov, Alexander Kolsanov, Tomislav Šmuc

## Abstract

Digital health technologies can reduce workloads, enhance information retention, and minimize errors. However, when poorly integrated or misaligned with clinical practice, they often introduce new risks and allow unprocessed information to accumulate, amplifying existing challenges. The core problem is that reasoning often stays implicit and unstructured when translating needs into digital solutions. Without a shared way to make it explicit, critical inputs are missed, unprocessed information builds up, and reasoning faults spread through the system. We present a minimal, non-redundant set of eight plain-language prompts that spans the full reasoning process, guards against common reasoning faults, and provides a shared structure for aligning perspectives across stakeholders. We tested the set in a clinical case study on the initial management of septic shock, where the prompts helped to turn unstructured clinical reasoning into clear inputs for system design and workflow adjustments. To interpret its impact, we built a mathematical model that captures how unprocessed information changes over time. Compared with standard practice, the prompts reduced peak backlog by 20.3%, cumulative backlog by 22.2%, and improved recovery time from *not reached* to five time steps. We share an open-source code to model backlog dynamics, enabling easy adaptation to other clinical settings through parameter calibration.

**Author summary:** In digital health, the biggest challenge is not the technology but translation. What clinicians need and what developers create often do not align, resulting in tools that fail to meet the demands of real-world care. One key reason is that clinicians, developers, and administrators approach the same problem from different perspectives, and their reasoning is rarely made explicit or shared in a structured way. Our solution is a minimal yet complete set of plain-language prompts. These prompts make reasoning clear, bring experts onto the same page, and help to avoid common thinking traps like missing critical details, focusing too narrowly, or hesitating to act. In a clinical case of septic shock management, the prompts kept essential information visible, helped to set priorities in real time, and produced inputs directly relevant to patient care. By making critical details more visible, the prompt set enabled faster responses and led to fewer delays in treatment, while maintaining reliability. This set is compact enough for quick adoption and effective enough to bring together the perspectives of diverse expert groups, making it useful not only in hospitals but also in public health and other complex care settings.

## Introduction

*A neonate is admitted to a Denver hospital. Born to a mother with a history of syphilis, the indication for treatment remains unclear due to language barriers and incomplete records. After consulting with infectious disease specialists, the physician prescribes benzathine penicillin G. The pharmacist, unfamiliar with neonatal dosing, misreads the standard dose as 500,000 units/kg instead of 50,000 units/kg. The handwritten “150,000U” order adds to the confusion. The “U” for units appears as an extra zero, resulting in a tenfold overdose. When the medication reaches the nurses, they hesitate, questioning the pain of five intramuscular injections for a newborn. Seeking an alternative, they check a drug reference book that does not explicitly warn against intravenous use. The nurses see “Benzathine” capitalized and placed on a separate line from “penicillin G*.*” They assume it is a brand name rather than a distinct formulation. The warning on the syringe “for intramuscular use only” is too small to read and goes unnoticed. The nurses know that only clear liquids are typically safe for intravenous use but believe some milky medications, like lipid-based drug products, can also be given intravenously. They proceed. After 1*.*8 ml is given, the baby becomes unresponsive and dies from a pulmonary embolism. An autopsy confirms the baby never had congenital syphilis. Three nurses are indicted, but expert testimony reveals over fifty system failures – misread orders, unclear references, and unchecked assumptions – turning small gaps into a fatal mistake. The public sees a clear-cut story of human error: they gave the wrong drug in the wrong way*.

*– The Denver medication error/criminal negligence case, 1998 [1]*

Since that tragedy healthcare has undergone significant changes through digital transformation, yet similar incidents continue to occur [2]. Across the world, national and institutional reporting systems [3], designed to capture clinical near misses and incidents [4–6], have documented countless recurring patterns. These events are rarely the result of a single error [7]. Instead, they typically arise from information gaps that result from shortcomings in how health systems are designed [8–10].

In an ideal world, healthcare practitioners would have immediate access to all the data needed to make informed decisions. In reality, the information they see – and what they notice within it – depends on their role, experience, current circumstances, and how their workflow is organized. Strict regulations meant to standardize how data is recorded and shared can limit flexibility, and existing infrastructure further shapes how healthcare systems function. But, system limitations are not the only issue. Human cognition narrows the flow of information even more. Our sensory memory briefly captures a wide range of inputs for only a fraction of a second. From there, only some of these inputs move into short-term memory, which typically retains them for less than half a minute. During that time, working memory manipulates this data before a portion of it is encoded into long-term memory, which can last from days to years.

These internal processes shape how we perceive events and handle incomplete information. When crucial details are lost, whether due to system constraints or human limitations [11–13], the result can be the very gaps illustrated in the opening Denver incident.

Human cognition, molded by evolutionary pressures, is not optimized for instantly handling large volumes of data. Instead, it simplifies, filters, and interprets information based on the inputs available at the moment [12, 14]. When information is scarce, the mind fills gaps with plausible but sometimes inaccurate assumptions [15–17]. When information is excessive, the limits of working memory [18–21] can cause cognitive overload [22–24]. In both cases, critical inputs remain unaddressed, creating a backlog of unprocessed information. Different reasoning faults contribute to this backlog in distinct ways. Omission prevents key data from entering the reasoning process [25], tunnel vision allows other relevant inputs to accumulate unresolved [26–29], clutter overwhelms limited processing capacity [30], and overconfidence lets incomplete or inaccurate information circulate unchecked [31]. This growing backlog not only slows reasoning and increases the risk of errors but also highlights the need for solutions specifically designed to reduce it.

Digital health tools, by design, aim to support this need by reducing the cognitive burden of managing complex information. When well integrated, they can ease workload [32, 33], help clinicians retain critical information [34–36], and reduce the risk of serious oversights [37, 38]. However, when poorly aligned with real-world workflows [39, 40], they may introduce new vulnerabilities [41, 42] and, in some cases, worsen the very problems they were intended to solve [43, 44].

Existing approaches to digital health design have focused on measuring cognitive burden [45–50] and recognizing fundamental differences between human and digital information processing [14, 36, 37]. Others have emphasized the importance of aligning how information is represented and understood across stakeholders [51–54]. However, gaps remain. Domain experts often struggle to fully articulate their reasoning, or they assume their needs have been clearly communicated when others have understood something entirely different. Without a structured way to externalize reasoning, each stakeholder brings a different frame of reference: clinicians focus on symptoms, risks, and urgency; developers on data models and workflow triggers; and administrators on efficiency and reporting metrics. As a result, the system fails to capture critical inputs in a consistent way, allowing a backlog of unprocessed information to accumulate and amplifying reasoning faults across the system.

This study presents a non-redundant, complete set of eight plain-language prompts. The prompts mitigate common reasoning faults in time-critical clinical situations and provide a shared framework for making reasoning explicit. We tested this approach in a clinical case study, using the prompts to generate clear inputs ready for translation into technical requirements or workflow improvements. We also developed a mathematical model that interprets how these prompts influenced the backlog of unprocessed information in the case study, making their impact easier to evaluate.

This study includes three main elements: the design of the minimal prompt set, a clinical case study demonstrating its practical application, and an interpretation of its effects on the accumulation of unprocessed information, highlighting reasoning bottlenecks. Figure 1 provides an overview of the full process, showing the underlying problems (left), the study design (center), and the impact (right).

**Fig 1.**
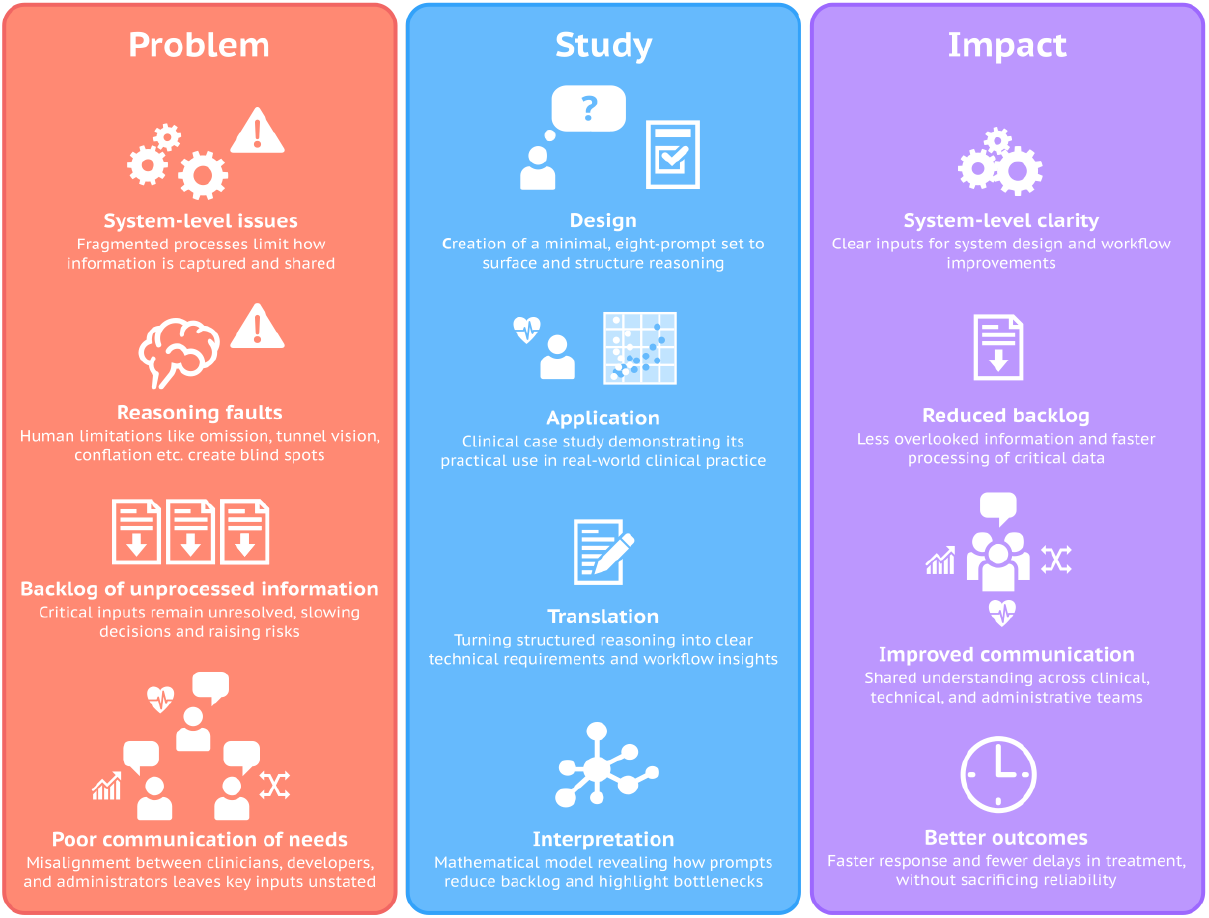
Overview of the study. The figure outlines the full process, starting with the underlying problems (left), moving through the study design (center), and ending with the expected impact (right). The approach targets system-level gaps, reasoning faults, and backlogs of unprocessed information by introducing a minimal eight-prompt set, testing it in a clinical case study, and interpreting its effects with a mathematical model. The resulting impact includes clearer inputs for design and operations, reduced backlog, better communication, and faster clinical responses without losing reliability.

## Results

### Prompt set

The prompt set is a compact collection of plain-language questions that guide the full reasoning process across four stages: Search, Shape, Check, and Act. Whereas condition-specific checklists in clinical practice are typically applied once a diagnosis has been established and treatment is underway, the prompts support reasoning in situations where information is still incomplete and uncertainty is high. Each prompt targets one of eight common reasoning faults – omission, tunnel vision, conflation, fragmentation, clutter, misprioritization, overconfidence, and paralysis – and the types of unprocessed information they create. This design makes reasoning gaps visible in real time and helps reduce the backlog of unprocessed information that slows responses and introduces delays.

Table 1 serves as a practical reference. For each prompt, it lists the guiding question, short instructions, and the type of backlog issue it helps resolve. Collectively, these components establish a structured way to make reasoning explicit. The underlying principles guiding the design of the prompt set are described in the Materials and methods section.

**Table 1.**
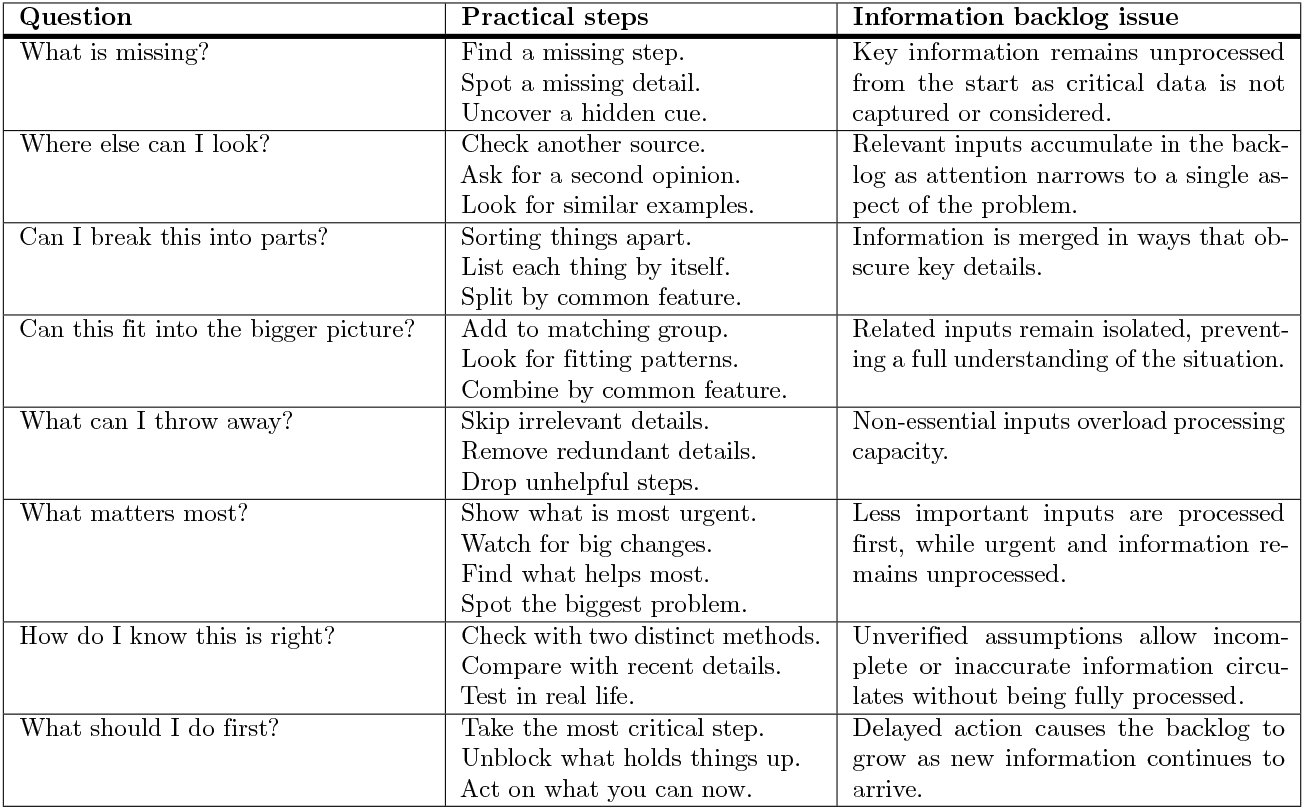
Prompt reference. Each prompt includes a guiding question, practical steps, and the backlog issue it addresses.

| Question | Practical steps | Information backlog issue |
| --- | --- | --- |
| What is missing? | Find a missing step.<br>Spot a missing detail.<br>Uncover a hidden cue. | Key information remains unprocessed from the start as critical data is not captured or considered. |
| Where else can I look? | Check another source.<br>Ask for a second opinion.<br>Look for similar examples. | Relevant inputs accumulate in the backlog as attention narrows to a single aspect of the problem. |
| Can I break this into parts? | Sorting things apart.<br>List each thing by itself.<br>Split by common feature. | Information is merged in ways that obscure key details. |
| Can this fit into the bigger picture? | Add to matching group.<br>Look for fitting patterns.<br>Combine by common feature. | Related inputs remain isolated, preventing a full understanding of the situation. |
| What can I throw away? | Skip irrelevant details.<br>Remove redundant details.<br>Drop unhelpful steps. | Non-essential inputs overload processing capacity. |
| What matters most? | Show what is most urgent.<br>Watch for big changes.<br>Find what helps most.<br>Spot the biggest problem. | Less important inputs are processed first, while urgent and information remains unprocessed. |
| How do I know this is right? | Check with two distinct methods.<br>Compare with recent details.<br>Test in real life. | Unverified assumptions allow incomplete or inaccurate information circulates without being fully processed. |
| What should I do first? | Take the most critical step.<br>Unblock what holds things up.<br>Act on what you can now. | Delayed action causes the backlog to grow as new information continues to arrive. |

By connecting reasoning faults to backlog issues, the prompts provide a systematic way to uncover system-level gaps. We next demonstrate their use in a clinical case study on septic shock management, showing how they helped identify these gaps and how structured reasoning can be translated into clear technical requirements and targeted workflow improvements.

### Clinical case study

#### Context

We tested the prompt set in a clinical case study of septic shock, chosen because of its rapid progression, high mortality, and the frequent need to manage incomplete or conflicting information. The patient profile reflects an initial assessment on arrival commonly encountered in clinical practice: a male, aged 60–70 years, with impaired consciousness, diabetes mellitus, and cardiac arrhythmia, admitted to the emergency department with signs of septic shock. In addition to confirming sepsis and identifying its source, the clinical team needed to rule out both diabetic coma and acute cardiac pathology. The case was structured around established sepsis criteria, with team involvement and documentation procedures described in the Materials and methods.

#### Application

Across five observation points, a team of clinicians involved in the case study applied the prompts to structure reasoning in real time. This process exposed system-level issues or recurring gaps that disrupted information flow and clinical judgment. At each step, clinicians first defined the clinical context in terms of the findings available at that moment, and then identified the recurring issues where missing or delayed information slowed responses. A complete account of the case, including clinical findings, prompt responses, and issues identified at each step, together with clinician-documented scenarios showing how these issues arise in routine practice, is provided in Supplementary Information (S1 Table, S2 Table). The Sankey diagram (Fig. 2) provides a visual summary of the case study, showing how prompts linked to issues across the five time steps.

**Fig 2.**
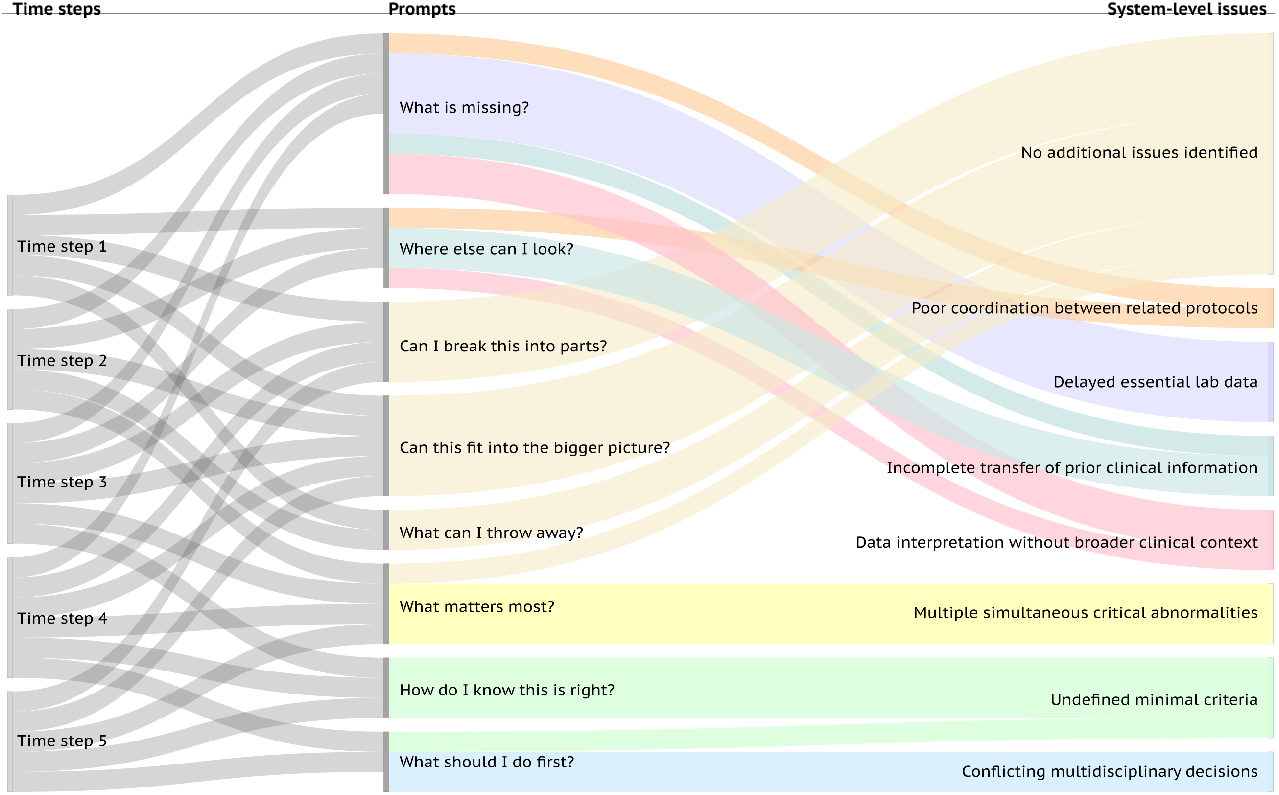
Application of the prompt set in a clinical case study on septic shock management. The Sankey diagram linking time steps (left), applied prompts (center), and identified system-level issues (right). At each time step, clinicians applied a subset of prompts to structure reasoning around available findings. This process revealed recurring system-level issues. The colors highlight distinct issue categories.

It indicates that at each step clinicians applied a subset of prompts, and across the complete sequence every prompt was used at least once. Distinct issue categories are highlighted by color.

Early in the process, *What is missing?* and *Where else can I look?* accounted for most issues (Steps 1–3), especially, delayed lab data, incomplete transfer of prior information, and poor coordination between related protocols. From Step 3 onward, *What matters most?* became dominant, surfacing multiple simultaneous critical abnormalities and showing how risks compound once partial data begins to arrive. The prompt *How do I know this is right?* repeatedly surfaced undefined minimal criteria, pointing to unclear confirmation rules across labs, imaging, and monitoring. *What should I do first?* was linked to conflicting multidisciplinary decisions at Steps 4–5, exposing friction within the team when urgent action was needed. Prompts that did not reveal new issues instead clarified available information, reducing ambiguity and keeping reasoning structured. To show how structured reasoning translates into clear inputs for system design and workflow improvements, we highlight an excerpt from the case study focusing on Steps 3–5 (Table 2), where recurring issues converge and intensify. These steps illustrate how clinical data accumulate over time, while the prompts expose system-level issues by structuring reasoning and reducing unprocessed information.

**Table 2.**
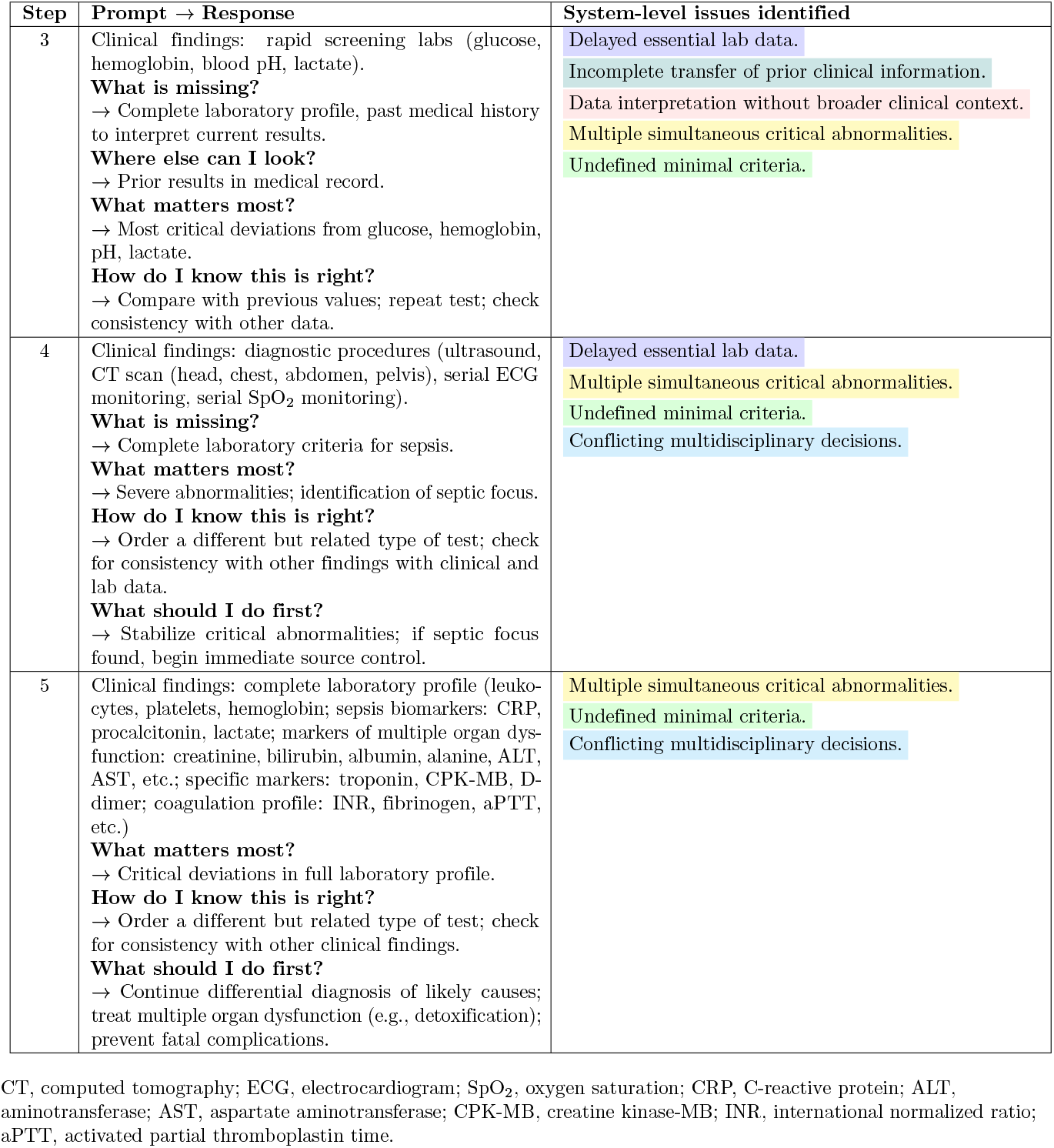
Excerpt from the septic shock case study. Each step shows the clinical findings available at that moment, the prompts used, and the system-level issues they revealed. The complete case study is provided in S1 Table.

| Step | Prompt → Response | System-level issues identified |
| --- | --- | --- |
| 3 | Clinical findings: rapid screening labs (glucose, hemoglobin, blood pH, lactate).<br>**What is missing?**<br>→ Complete laboratory profile, past medical history to interpret current results.<br>**Where else can I look?**<br>→ Prior results in medical record.<br>**What matters most?**<br>→ Most critical deviations from glucose, hemoglobin, pH, lactate.<br>**How do I know this is right?**<br>→ Compare with previous values; repeat test; check consistency with other data. | Delayed essential lab data.<br>Incomplete transfer of prior clinical information.<br>Data interpretation without broader clinical context.<br>Multiple simultaneous critical abnormalities.<br>Undefined minimal criteria. |
| 4 | Clinical findings: diagnostic procedures (ultrasound, CT scan (head, chest, abdomen, pelvis), serial ECG monitoring, serial SpO<sub>2</sub> monitoring).<br>**What is missing?**<br>→ Complete laboratory criteria for sepsis.<br>**What matters most?**<br>→ Severe abnormalities; identification of septic focus.<br>**How do I know this is right?**<br>→ Order a different but related type of test; check for consistency with other findings with clinical and lab data.<br>**What should I do first?**<br>→ Stabilize critical abnormalities; if septic focus found, begin immediate source control. | Delayed essential lab data.<br>Multiple simultaneous critical abnormalities.<br>Undefined minimal criteria.<br>Conflicting multidisciplinary decisions. |
| 5 | Clinical findings: complete laboratory profile (leukocytes, platelets, hemoglobin; sepsis biomarkers: CRP, procalcitonin, lactate; markers of multiple organ dysfunction: creatinine, bilirubin, albumin, alanine, ALT, AST, etc.; specific markers: troponin, CPK-MB, D-dimer; coagulation profile: INR, fibrinogen, aPTT, etc.)<br>**What matters most?**<br>→ Critical deviations in full laboratory profile.<br>**How do I know this is right?**<br>→ Order a different but related type of test; check for consistency with other clinical findings.<br>**What should I do first?**<br>→ Continue differential diagnosis of likely causes; treat multiple organ dysfunction (e.g., detoxification); prevent fatal complications. | Multiple simultaneous critical abnormalities.<br>Undefined minimal criteria.<br>Conflicting multidisciplinary decisions. |

CT, computed tomography; ECG, electrocardiogram; SpO<sub>2</sub>, oxygen saturation; CRP, C-reactive protein; ALT, aminotransferase; AST, aspartate aminotransferase; CPK-MB, creatine kinase-MB; INR, international normalized ratio; aPTT, activated partial thromboplastin time.

#### Translation

The issues identified in Steps 3–5 were translated into four event-driven and state-driven requirements, summarized in the blocks below: keeping missing data visible until resolved, summarizing critical abnormalities by organ system, maintaining a shared record of urgent priorities across teams, and making diagnostic uncertainty explicit until standardized criteria are available.

**Requirement 1 (Event-driven)**

**When** results or history are missing, **the system shall** show them clearly and mark decisions as temporary.

*This prevents premature decisions from being treated as final*.

###### System-level issue addressed

Delayed essential lab data; incomplete transfer of prior clinical information.

**Requirement 2 (Event-driven)**

**When** new critical findings appear, **the system shall** summarize them by organ system to show combined risk.

*This supports recognition of compounding risks*.

###### System-level issue addressed

Multiple simultaneous critical abnormalities.

**Requirement 3 (State-driven)**

**While** urgent priorities are being managed, **the system shall** keep a shared, updated list across teams.

*This ensures all teams act on the same priorities*.

###### System-level issue addressed

Conflicting multidisciplinary decisions.

**Requirement 4 (State-driven)**

**When** diagnostic criteria are unclear, **the system shall** highlight the uncertainty and align with standardized rules once available.

*This makes uncertainty explicit until clear rules are applicable*.

###### System-level issue addressed

Undefined minimal criteria.

These requirements illustrate how the prompts made critical gaps and risks more visible. In the next section, we model how the prompts affects the backlog of unprocessed information.

### Backlog dynamics

#### Mathematical model

Most existing models in healthcare focus on patient flow, task queues, or staff capacity. Our model instead captures the backlog of unprocessed information and how reasoning prompts shape it. By design, the prompt set aims to reduce the likelihood that crucial information will remain unprocessed. Building on this rationale, we developed a mathematical model to quantify how the prompts impact the backlog in clinical settings.

Let *t* = 0, 1, …, *n* denote the discrete observation steps, where *n* is the final step in the observation period. The interval between two consecutive steps is given by Δ*t*. These intervals do not need to be evenly spaced, so their length may vary throughout the observation period. Let *I*_*t*_ be the quantity of unprocessed information at time *t*. The discrete-time dynamics are given by

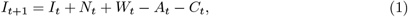

where

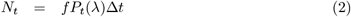

is the new information arriving in the interval from *t* to *t* + 1, with *f >* 0 a scaling parameter and *P*_*t*_(*λ*) the Poisson probability mass function with mean *λ*;

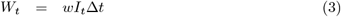

is the noise term, with *w* ≥ 0 quantifying the intensity of additive noise proportional to the current backlog (that is, the unprocessed information at time *t*). The parameter *w* may vary over time due to organizational factors such as infrastructure and information flow within a medical facility, but is treated as constant to clarify the analysis of prompt effects;

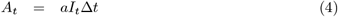

is the amount of information processed by the clinical team during the interval Δ*t*, where *a >* 0 denotes the intrinsic processing rate. The parameter *a* may, in general, be time dependent, reflecting factors such as fatigue, staffing, or patient load, but is held constant here to isolate the effects of the prompts;

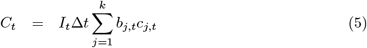

represents the combined impact of all selected prompts on the backlog at step *t*. Here, *j* = 1, …, *k* indexes the set of available prompts, where *b*_*j,t*_ ∈ 0, 1 indicates whether the *j*-th prompt is applied at time *t* (1 if applied, 0 otherwise), and *c*_*j,t*_ describes the immediate fractional effect of the *j*-th prompt on the backlog.

A positive value *c*_*j,t*_ *>* 0 indicates a reduction in unprocessed information, while *c*_*j,t*_ *<* 0 reflects an increase. This behavior describes the expected impact of prompts. For example, *What is missing?* often uncovers gaps (e.g., pending labs, no history available). Making these gaps explicit temporarily enlarges the backlog (*c*_*j,t*_ *<* 0).

However, this effect is constructive, since without the prompt the gaps would remain hidden, leaving the system unreliable. The prompts therefore do not always shrink the backlog immediately. Instead, they reshape it by exposing hidden issues that can then be addressed. In addition, the prompts interact dynamically. Resolving one issue may give rise to another. This transformation of issues is represented in the model by the time-varying values of *c*_*j,t*_.

#### Steady-state behavior

Steady-state analysis determines whether unprocessed information is eventually eliminated or persists in the system. Expanding Eq (1) with the definitions in Eq (2)–Eq (5) gives

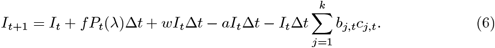

To find the equilibrium, set *I*_*t*+1_ = *I*_*t*_ = *I*^*^, where *I*^*^ is the steady-state value.

Substituting this into Eq (6) above yields

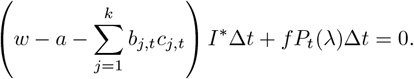

As *t* → ∞, the arrival rate of new information *P*_*t*_(*λ*) approaches zero, so *I*^*^ = 0. Thus, the unique equilibrium corresponds to complete elimination of unprocessed information.

#### Stability analysis

Stability analysis is crucial for understanding whether the system returns to equilibrium after small disturbances. Consider a small perturbation *δI*_*t*_ from equilibrium. For large *t*, the *f P*_*t*_(*λ*) term can be omitted, since it vanishes as the process concludes. The linearized update Eq (6) for the perturbation becomes

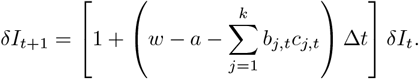

Stability holds when the absolute value of the multiplier is less than one, as this guarantees that small deviations from the equilibrium decay with each time step:

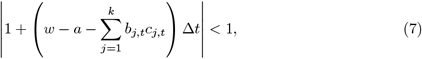

which makes all perturbations decay over time. For small enough time steps Δ*t*, Eq (7) reduces to

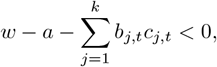

thereby ensuring *I*^*^ = 0 is a stable equilibrium.

#### Minimum and maximum values

The minimum and maximum values of *I*_*t*_ (Eq (6)) provide additional insight into the system behavior, as these extremes indicate the potential range of unprocessed information during the observation period. Understanding these values is crucial for optimizing resource allocation: if the maximum value of *I*_*t*_ is too high, it may indicate periods when the team is overloaded and additional personnel or capacity are required to process the incoming information effectively. The minimum is *I*_*t*_ = 0, since a negative backlog is not possible. The maximum depends on the initial value, the time dependence of *P*_*t*_(*λ*), and the specific values of *a, w*, as well as the selection of *b*_*j,t*_ and *c*_*j,t*_. Typically, *I*_*t*_ increases as new information arrives, reaches a maximum near the peak of *P*_*t*_(*λ*), and decreases as arrivals decline and processing dominates. The time step *t*^*^ at which *I*_*t*_ attains its maximum can be found by setting *I*_*t*+1_ − *I*_*t*_ = 0 in Eq (6):

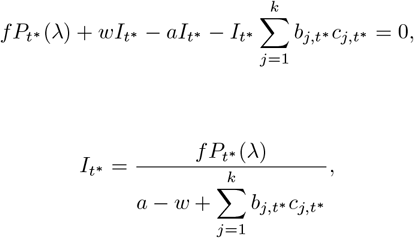

where *t*^*^ typically coincides with or near the peak of 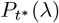.

By selecting parameter values *a, w, b*_*j,t*_, and *c*_*j,t*_ appropriate to the clinical context, the discrete-time model presented here can be used to assess how effectively the prompt set impacts unprocessed information. Details on related models, model design, and the selection of parameters (*c, a, λ*) are provided in the Materials and methods section.

### Interpretation

We applied the model to the septic shock case study to interpret the impact of the prompt set on the backlog of unprocessed information. The parameters were taken directly from the case study, reflecting the hospital setting, clinical team, infrastructure, and workflow.

Figure 3 shows the simulated backlog *I*_*t*_ (Eq. 1) across time steps for two scenarios: the red dashed line represents performance without prompts, and the blue solid line shows performance with the prompt set. With prompts, the backlog reached a lower maximum and declined faster than in the baseline. The simulation indicates that the maximum backlog, the peak backlog, was reduced by 20.3% with prompts. The total backlog accumulated across all steps, the cumulative backlog, was reduced by 22.2%. The recovery, defined as the first step when the backlog fell to one unit or less, occurred at step five with prompts but was not reached under the baseline scenario. Details of parameter selection and metric definitions are provided in Materials and methods. The implementation of the backlog model, along with parameter estimates and key metrics, is provided in S1 File.

**Fig 3.**
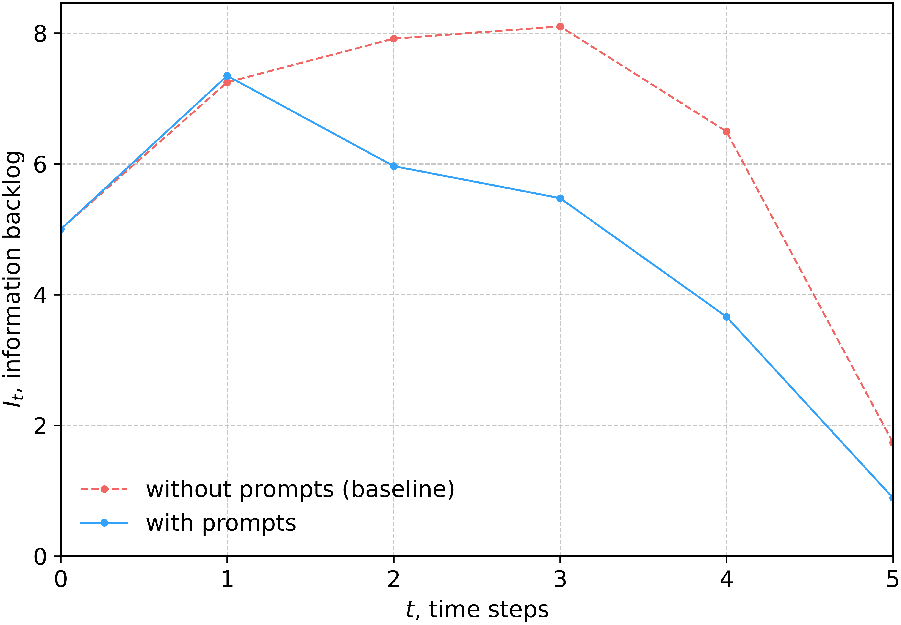
Impact of prompts on the backlog of unprocessed information. The simulated backlog *I*_*t*_ (Eq. (1)) for scenarios with and without prompts. With prompts, the peak backlog was 7.12 at *t* = 1; without prompts, 8.93 at *t* = 2 (20.3% reduction). With prompts, the cumulative backlog over the observation period was 22.2% lower, and recovery, defined as the first time the backlog falls to 1 unit or less, occurred at *t* = 5; without prompts, recovery was not reached.

These improvements have direct practical implications. Lower peak backlog reduced the risk of overlooking critical findings during high information load. Reduced cumulative backlog meant more data were processed in time, with essential information integrated rather than left unresolved. Faster recovery helped the team regain situational clarity earlier, supporting clearer inputs for system design, better communication across the team, and quicker, more reliable responses with fewer delays in treatment.

## Discussion

### Lessons learned

This study offers several practical insights and lessons learned. A key one was the value of plain-language prompts for structuring reasoning. Clinicians found that the prompts acted like ad-hoc checklists in situations of high uncertainty and limited time. Unlike fixed checklists, which assume the right pathway is already known, the prompts enriched the available information so that clinicians could decide which checklist or framework to apply. By breaking reasoning into simple steps, the prompts reduced ambiguity, surfaced overlooked issues, and kept critical information visible without adding cognitive burden.

Another lesson is that the prompts helped make system-level gaps visible. By tracing where information was missing, delayed, or poorly shared, clinicians could see patterns that pointed to workflow problems rather than individual oversights. This shifted attention from personal error to structural issues, providing clearer input for improving communication and digital support systems.

### Limitations

Several limitations should also be noted. The case study focused on a single condition, sepsis, in one hospital setting, which may limit direct generalizability. Second, the mathematical model was deliberately simplified to isolate the impact of prompts, and therefore does not capture the full complexity of team behavior, uncertainty, or evolving patient conditions. Third, the estimates for the key model parameters (*c, a, λ*) relied on clinician judgment, introducing subjectivity.

### Future directions

This work points to several directions for further development. The prompt set can be extended beyond point-of-care use by linking it to low-code and AI-assisted development, where clear requirements are essential for building responsible and clinically relevant digital tools.

Prompt mapping also has potential as an audit method. By comparing how prompts are used across cases, departments, or before and after digital interventions, hospitals could track shifts in workflow, identify which gaps are resolved, and detect new vulnerabilities. This could be particularly valuable in regulatory contexts, where evidence is needed to demonstrate that critical safety checks remain visible and no decision points are overlooked. Such audits could strengthen compliance reviews, target improvements, and reduce risks before widespread deployment.

Further testing is needed to evaluate generalizability. Future studies should extend the approach to other acute conditions such as trauma, stroke, and cardiac arrest, and validate the backlog model with real-time data across multiple hospitals. Beyond backlog dynamics, research should also assess how prompts influence teamwork, diagnostic accuracy, and patient outcomes. Finally, adaptive prompt delivery, such as context-sensitive suggestions integrated into electronic health records or dashboards, could further reduce cognitive load while preserving clarity.

## Materials and methods

### Prompt set design

The prompt set was designed to be minimal yet complete, covering the full reasoning process without redundancy. The design followed three principles: (1) each stage of reasoning is represented, (2) every prompt addresses a distinct reasoning fault documented in the literature [25–31], and (3) each fault is linked to a type of unprocessed information that slows clinical responses.

The reasoning process was structured into four stages: Search, Shape, Check, and Act. These reflect common patterns in clinical reasoning, moving from gathering inputs to organizing them, checking priorities and assumptions, and deciding on actions. Each stage was linked to specific reasoning faults and the types of unprocessed information (Table 3).

**Table 3.** Prompt design across reasoning stages. Each stage connects guiding questions with reasoning faults and types of unprocessed information.

| Stage | Reasoning fault | Type of information | Guiding question | Practical steps |
| --- | --- | --- | --- | --- |
| Search | Omission | Uncaptured inputs | What is missing? | Find a missing step.<br>Spot a missing detail.<br>Uncover a hidden cue. |
|  | Tunnel vision | Piling inputs | Where else can I look? | Check another source.<br>Ask for a second opinion.<br>Look for similar examples. |
| Shape | Conflation | Merged inputs | Can I break this into parts? | Sorting things apart.<br>List each thing by itself.<br>Split by common feature. |
|  | Fragmentation | Isolated inputs | Can this fit into the bigger picture? | Add to matching group.<br>Look for fitting patterns.<br>Combine by common feature. |
|  | Clutter | Irrelevant inputs | What can I throw away? | Skip irrelevant details.<br>Remove redundant details.<br>Drop unhelpful steps. |
| Check | Misprioritization | Deferred urgent inputs | What matters most? | Show what is most urgent.<br>Watch for big changes.<br>Find what helps most.<br>Spot the biggest problem. |
|  | Overconfidence | Unchecked inputs | How do I know this is right? | Check with two distinct methods.<br>Compare with recent details.<br>Test in real life. |
| Act | Paralysis | Delayed inputs | What should I do first? | Take the most critical step.<br>Unblock what holds things up.<br>Act on what you can now. |

Each stage–fault link was expressed as a plain-language guiding question, paired with three to four short steps to make it usable in practice. The steps drew on clinical practice, where clinicians described how they reason under pressure, such as checking another source or asking for a second opinion. They were also informed by literature on cognitive strategies, aligning with heuristics like listing, grouping, or cross-checking to address reasoning faults. Finally, they were written in plain, action-oriented language so they could be applied in real clinical settings.

To keep the set both minimal and complete, prompts were mapped onto a stage–fault matrix to ensure that each reasoning fault was addressed in the right stage. We then tested the set by removing prompts one by one. If removing a prompt left a fault uncovered or made another prompt carry overlapping functions, the prompt was kept. This process confirmed that all eight prompts were necessary to ensure complete coverage without overlap. The resulting set balances completeness with conciseness, aligning with Miller’s 7 *±* 2 principle on limitation of working memory [18] to remain practical under time pressure.

### Case study setup

The case study was structured around the qSOFA criteria [55] to ensure clinical relevance and alignment with established diagnostic frameworks. A team of clinicians experienced in emergency care and sepsis management estimated that the course of events from initial assessment on arrival to achieving a sufficient clinical picture for diagnosis corresponded to approximately five time points and 30 minutes in real practice.

At each time point, the clinicians documented the findings available at that moment and worked through the prompts in the order provided in the prompt reference

(Table 1), skipping those not applicable. This structured reasoning process highlighted repeating prompts, which were grouped according to problems in the information flow. From these patterns, the clinicians identified recurring issues that reflected system-level gaps within the medical facility. For each issue, they also provided clinical scenarios from their routine practice where such problems typically arise.

Clinicians organized their work using a structured layout that matched their reasoning process and made the case easier to document (S2 File). Each time point was placed at the top of a column, followed by the clinical findings available at that stage. Beneath, clinicians recorded their responses to the prompts, highlighting each stage of reasoning with a distinct color: white for finding missing information, blue for organizing and simplifying what is known, black for checking priorities and assumptions, and red for deciding what to do first. At the bottom, they reserved a separate space to list the system-level issues that surfaced during the process and show which prompt had triggered them. This layout allowed the team to track how information accumulated, how reasoning evolved, and how recurring issues emerged across steps.

The identified issues were reviewed with hospital administrators, who assessed their relevance to existing workflows and potential digital interventions. Developers then formalized the prompt responses and issues into structured software requirements using the Easy Approach to Requirements Syntax (EARS) [56, 57], ensuring consistency and reducing ambiguity, omission, and complexity.

### Model parameters and metrics

The backlog model parameters were derived directly from the septic shock case study described above. This case, conducted in a hospital setting, reflected the characteristics of its clinical team, infrastructure, and workflow. Each parameter was selected to mirror this setting, ensuring that the model captured both the pace of new clinical inputs and the team’s capacity to process them (Table 4).

**Table 4.** Scalar parameters. Model parameters of the backlog model derived from the septic shock case study.

| Parameter | Description | Value |
| --- | --- | --- |
| $n$ | Number of time steps | 5 |
| $k$ | Number of prompts | 8 |
| $a$ | Fraction of backlog resolved per step | 0.5 |
| $w$ | Noise scaling coefficient | 0.15 |
| $\lambda$ | Rate of incoming information | 1.2 |
| $f$ | Scale factor for incoming information | 15 |
| $I_0$ | Initial backlog | 5 |

The arrival of new information was modeled as a Poisson process. The rate parameter *λ* was set to 1.2 to reflect the average number of new findings observed during the early phase of the case, when laboratory and monitoring results began to accumulate. The scale factor *f* was set to 15 to match the typical volume of information arriving per step. The initial backlog *I*_0_ was set to 5, corresponding to the number of patient parameters documented at admission before additional findings began to accumulate.

The team’s capacity to handle information was described by the processing rate coefficient *a*, set to 0.5 to represent the ability to resolve about half of the backlog per step. Since one step corresponded to 5–6 minutes (five steps over 30 minutes), this rate reflects a realistic pace for emergency care. The noise coefficient *w* was fixed at 0.15, representing the effect of unpredictable disturbances such as delayed lab results or communication bottlenecks.

Prompt use was represented by a schedule *b*_*j,t*_ (Table 5) and effect values *c*_*j,t*_ (Table 6). These inputs captured when prompts were applied in practice and how they impacted backlog dynamics.

**Table 5.**
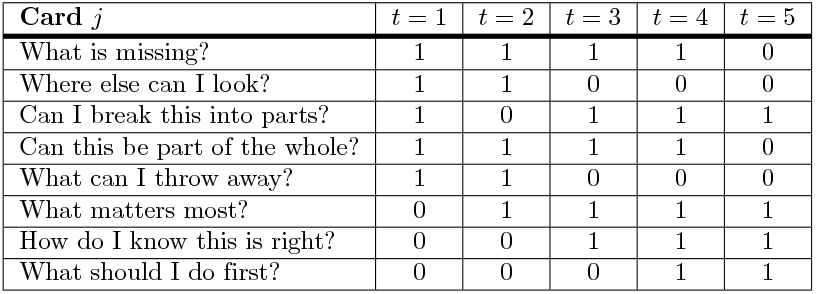
Prompt schedule. Schedule showing which prompts were applied at each observation step.

| Card $j$ | $t = 1$ | $t = 2$ | $t = 3$ | $t = 4$ | $t = 5$ |
| --- | --- | --- | --- | --- | --- |
| What is missing? | 1 | 1 | 1 | 1 | 0 |
| Where else can I look? | 1 | 1 | 0 | 0 | 0 |
| Can I break this into parts? | 1 | 0 | 1 | 1 | 1 |
| Can this be part of the whole? | 1 | 1 | 1 | 1 | 0 |
| What can I throw away? | 1 | 1 | 0 | 0 | 0 |
| What matters most? | 0 | 1 | 1 | 1 | 1 |
| How do I know this is right? | 0 | 0 | 1 | 1 | 1 |
| What should I do first? | 0 | 0 | 0 | 1 | 1 |

**Table 6.**
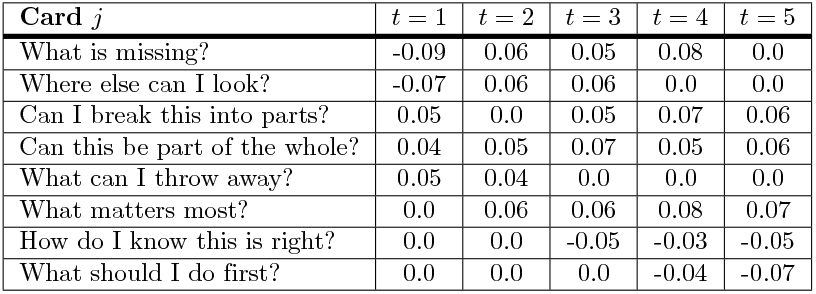
Prompt effects. Estimated impact of each prompt on the backlog of unprocessed information at each observation step.

Using this parameterization, we simulated backlog dynamics for two scenarios: with prompts and without prompts. To interpret the impact of the prompt set, model performance was assessed with three complementary metrics, each capturing a different aspect of backlog behavior:

The peak backlog, defined as the highest backlog observed across the observation period,

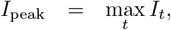

the total backlog accumulated across all steps,

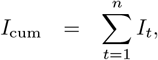

and the recovery time, defined as the first step when the backlog falls to one unit or less:

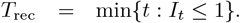

These metrics enabled comparison of the scenarios with and without prompts by capturing backlog intensity, total accumulation, and recovery time.

## Data Availability

All relevant data are within the manuscript and its Supporting Information files.

## Supporting information

**S1 Table. Prompt set in the initial management of septic shock**. A documented case study illustrates how prompts were applied to identify system-level issues affecting information flow and clinical judgment. Prompts not applicable at a given step were skipped.

**S2 Table. Clinician-reported scenarios illustrating system-level issues**. Documented scenarios from routine clinical practice show how each issue can arise in real-world settings.

**S1 File. Backlog dynamics notebook**. A Jupyter notebook provides parameter estimates from the case study, model implementation, and key metrics to compare system performance with and without prompts.

**S2 File. Structured case study documentation**. A clinician-documented case study shows how prompts were applied across time points, with clinical findings, responses, and system-level issues recorded in an intuitive layout. Reasoning stages are highlighted in color, and issues are linked directly to the prompts that revealed them.

## Acknowledgments

We thank all participants for sharing their perspectives during the pilot program, which contributed to refining the prompt set. We are also grateful to Dr. Mate Lerga from the Clinical Hospital Center, Rijeka, Croatia for his assistance in developing the clinician-reported scenarios used in the case study.

---

**S1 Table.**
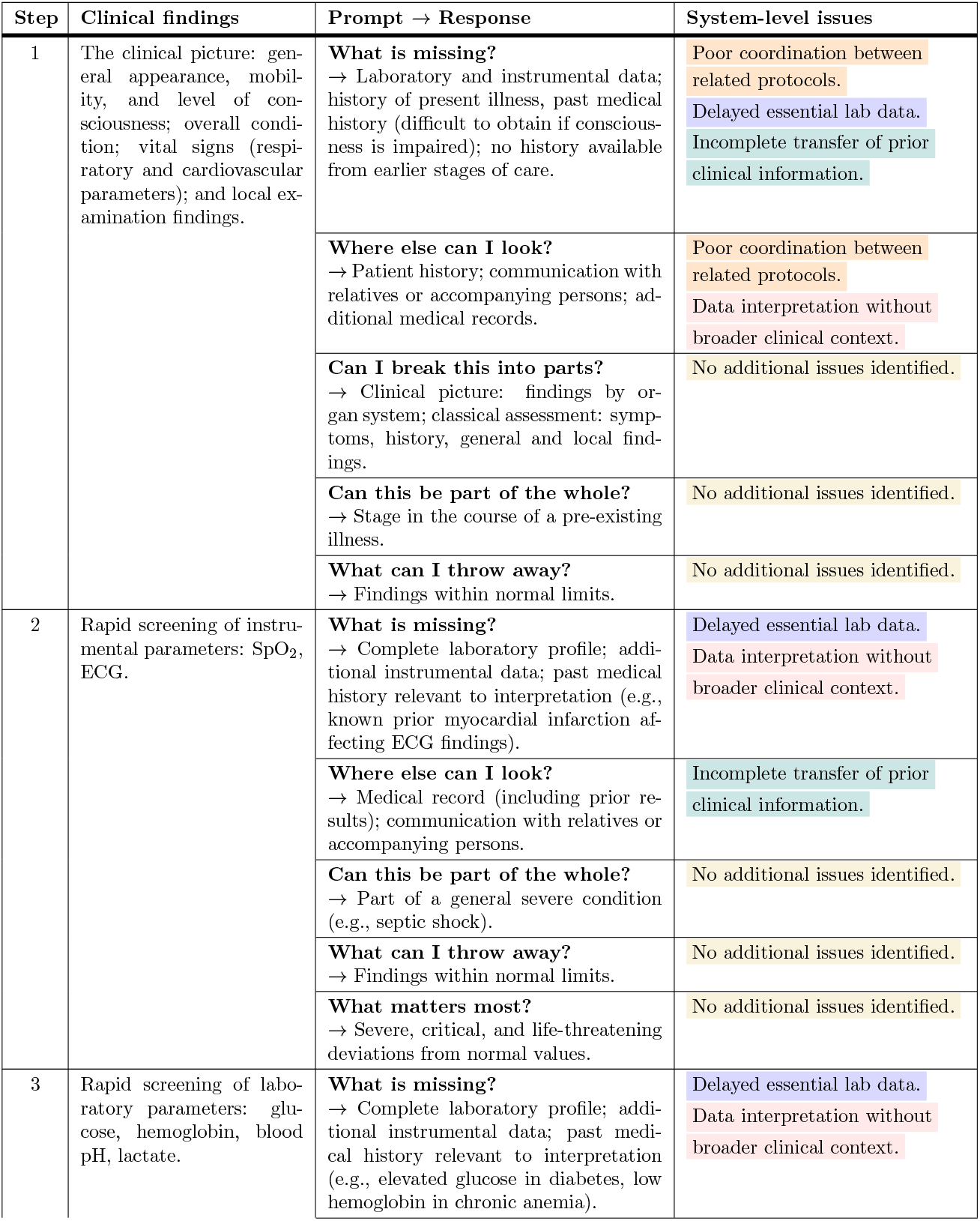
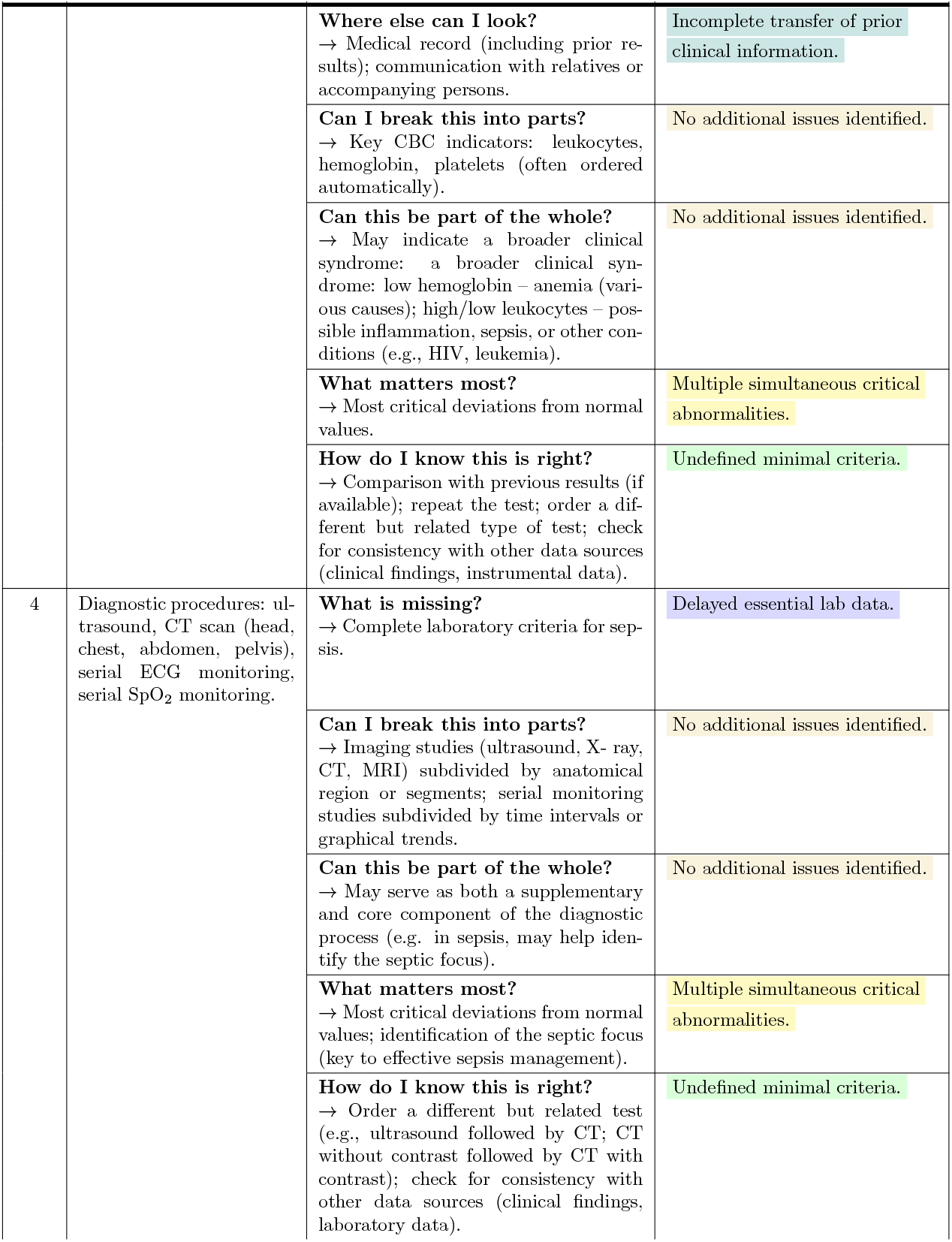
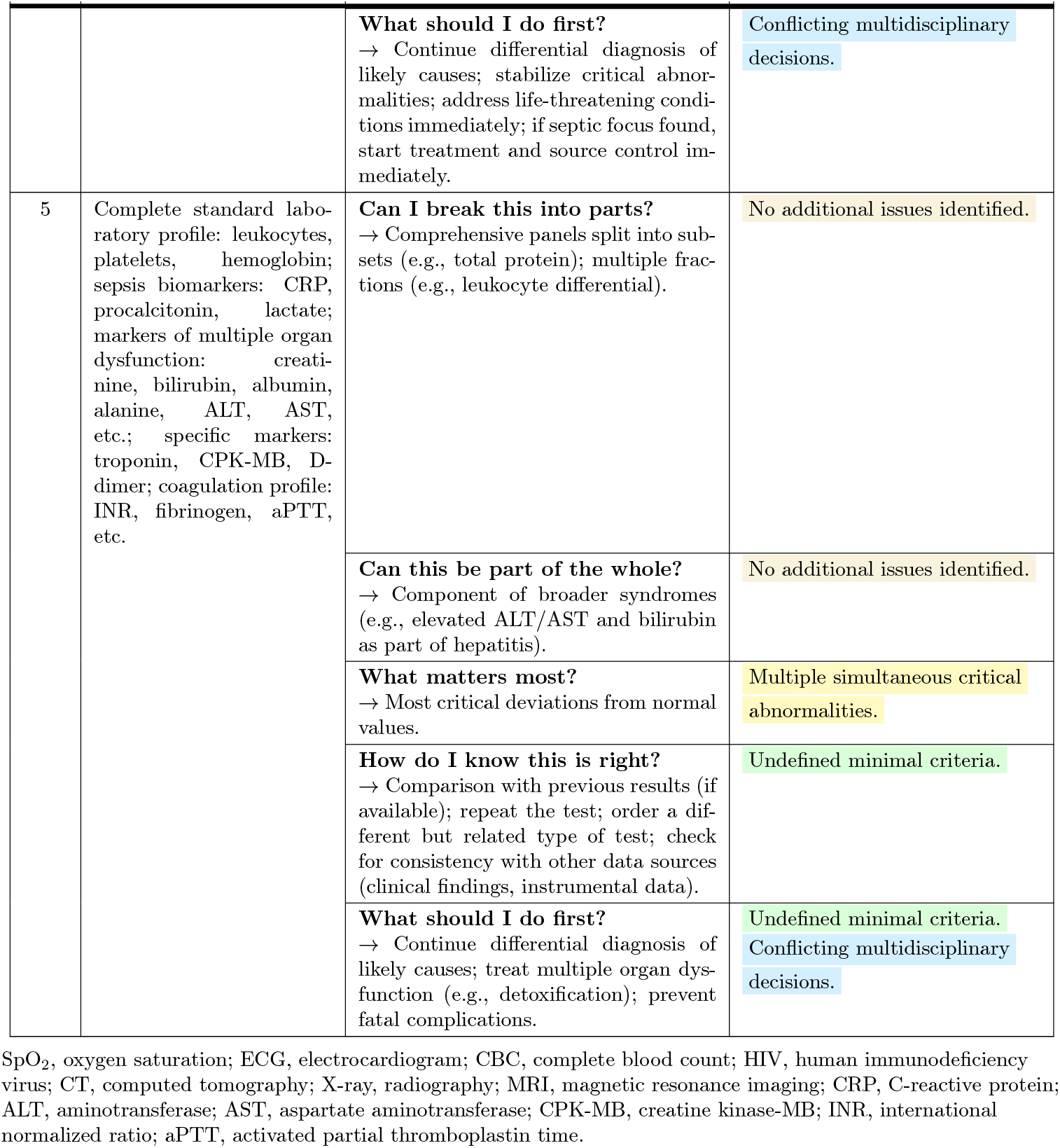
Prompt set in the initial management of septic shock. Structured case study showing how prompts were applied to identify system-level issues affecting information flow and clinical judgment. Prompts not applicable at a given step were skipped.

**S2 Table.** Clinician-reported scenarios illustrating system-level issues. Scenarios from routine clinical practice showing how each issue may arise in real-world settings.

| System-level issues | Clinical scenarios |
| --- | --- |
| Poor coordination between related protocols | In a patient with insulin-dependent diabetes, insulin administration and meal times were misaligned, increasing the risk of hypoglycemia. Clinicians informally adjusted meal schedules to align with insulin injections, ensuring patient safety. |
| Undefined minimal criteria | Clinicians delayed treatment due to uncertainty about the minimal clinical criteria required to confirm the diagnosis and justify immediate action. |
| Delayed essential lab data | Blood cultures typically require several days, forcing clinicians to make initial decisions based on incomplete information. |
| Multiple simultaneous critical abnormalities | In sepsis, multiple abnormalities often appear at once, indicating multi-organ dysfunction. Patients commonly present with elevated leukocytes, CRP, procalcitonin, ALT, AST, creatinine, bilirubin, and glucose, along with reduced hemoglobin and platelets. At the same time, life-threatening conditions such as severe hypoxia, hypotension, rapid heart rate, and decreased urine output arise. Clinicians must rapidly prioritize managing immediate threats, like low oxygen levels and shock, before addressing other abnormalities and treating the underlying infection. |
| Data interpretation without clinical context | Initially, a CRP level of 100 mg/L appeared concerning because it exceeded normal values. However, comparing it to a previous value of 200 mg/L revealed significant improvement. |
| Conflicting multidisciplinary decisions | A surgeon ordered an intestinal X-ray using barium sulfate to check for bowel obstruction, but the barium created imaging artifacts that interfered with a later CT scan.<br>In another case, a surgeon recommended diagnostic laparoscopy for a patient with unclear symptoms, but anesthesiologists opposed the procedure due to concerns about potential complications from intubation and difficulties restoring natural breathing afterward. |
| Incomplete transfer of prior clinical information | During ambulance transport from an outpatient facility to the hospital, the medical record of a patient with impaired consciousness was lost, resulting in the absence of crucial information about chronic illnesses and current medications. |

CRP, C-reactive protein; ALT, alanine transaminase; AST, aspartate transaminase; CT, computed tomography.

## Structured case study documentation

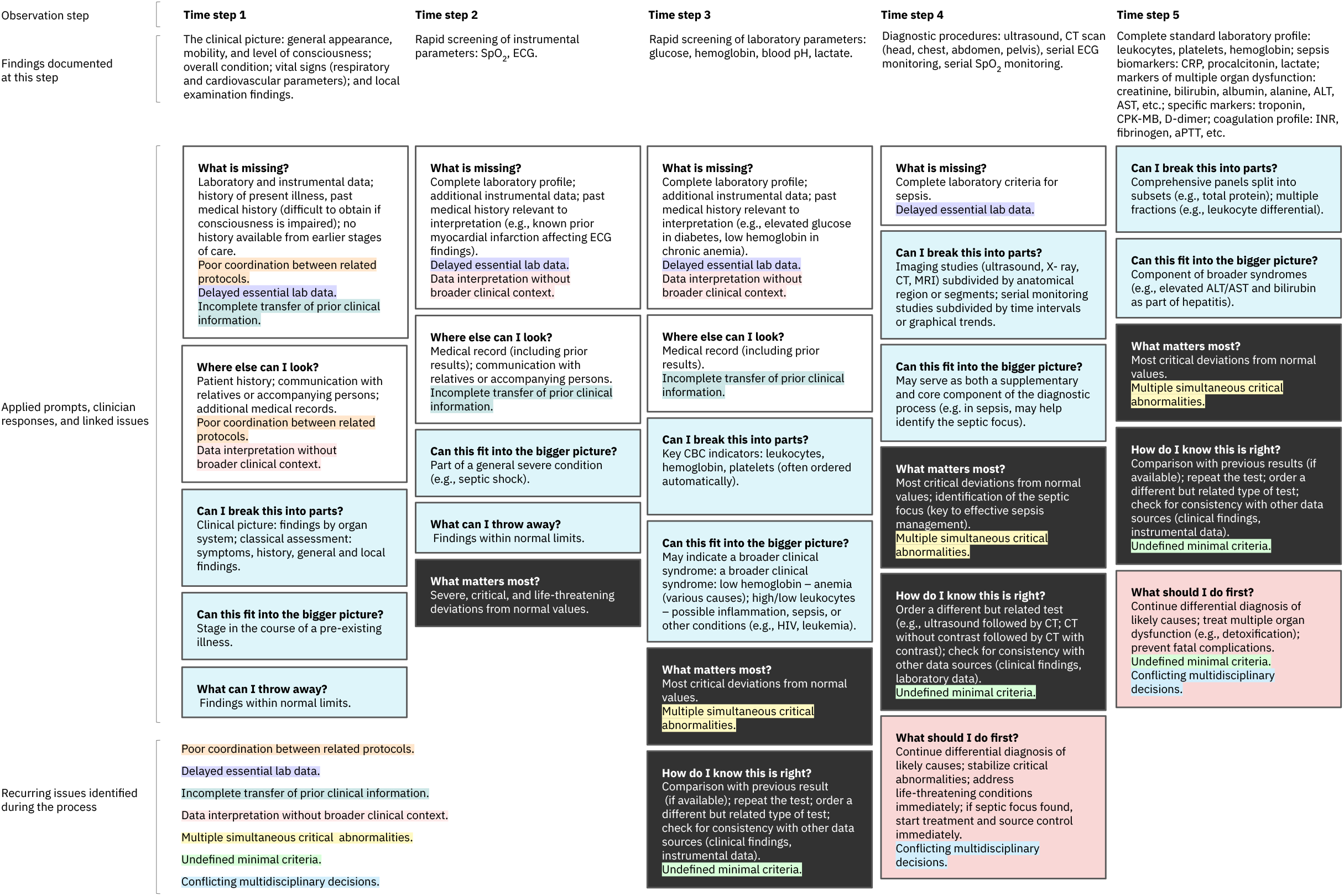

